# Population-based estimates of the prevalence of children with congenital heart disease and associated comorbidities

**DOI:** 10.1101/2023.06.29.23292065

**Authors:** Devin M. Parker, Meagan E. Stabler, Todd A MacKenzie, Meghan S. Zimmerman, Xun Shi, Allen D. Everett, Emily M. Bucholz, Jeremiah R. Brown

## Abstract

**Background:** Congenital heart defects (CHD) are the most common birth defects and are estimated to affect almost 1% of births annually in the US. CHDs are the leading cause of infant mortality associated with birth defects and can result in chronic disability, morbidity and increased healthcare costs. Most CHD prevalence estimates rely on data from population-based birth defects surveillance systems and these estimates are inconsistent due to varied definitions. It is therefore important to derive high-quality, population-based estimates of the prevalence of CHD to help care for this vulnerable population.

**Methods:** We performed a descriptive, retrospective 8-year analysis using all payer claims data (APCD) from Colorado (CO) from 2012 - 2019. Children with CHD were identified by applying ICD-9 and ICD-10 diagnoses codes from the American Heart Association-American College of Cardiology (AHA-ACC) harmonized cardiac codes. We included children with CHD <18 years of age who resided in CO, had a documented zip code, and had at least one healthcare claim. CHD type was categorized as simple, moderately complex and complex disease. Association with comorbid conditions and genetic diagnoses were analyzed using chi-square test. We used direct standardization to calculate adjusted prevalence rates, controlling for age, sex, primary insurance provider and urban-rural residence.

**Results:** We identified 1,566,328 children receiving care in CO from 2012 – 2019. Of those, 30,512 children had at least one CHD diagnosis, comprising 1.9% (95% CI: 1.93 – 1.97) of the pediatric population. Over half of the children with CHD also had at least one complex chronic condition and 11% had a genetic diagnosis.

**Conclusion:** The current study is the first population-level analysis of pediatric CHD in the US. Our study found a higher CHD prevalence than in previously reported studies and a high comorbidity burden. These findings can inform initiatives to improve screening, treatment and planning of care for this complex population.

## INTRODUCTION

Congenital heart defects (CHD) are the most common birth defects and are estimated to affect almost 1% of births annually in the US.^1, 2^ CHDs are the leading cause of infant mortality associated with birth defects and can result in chronic disability, morbidity and increased healthcare costs.^3^

Previous prevalence estimates of children with CHD have been inconsistent due to varied definitions, included only a subset of CHD diagnoses, were geographically limited and were generated from a small number of sources.^3, 4^ Many frequently cited prevalence estimates were derived from birth defect registries, where a diagnosis was made in a regional pediatric cardiology center and relied on patient referrals. Studies that actively screen for cases of CHD by echocardiography are not done routinely.^4, 5^ A seminal report by Hoffman and colleagues in 1965 estimated prevalence of structural heart defects at birth in 6-11 cases per 1,000 births. While informative, their study excluded the most common CHD diagnoses - ventricular and atrial septal defects.^6, 7^ Hoffman et al. concluded that much of the observed variability across studies reflected differences in case ascertainment.^3, 6^

Another important source of variation is the nomenclature used in CHD prevalence studies. Disparate approaches have been used to identify CHD, including definitions specifically developed for congenital registries or screening programs and studies limited to CHD requiring surgery. The most widely cited prevalence estimates were generated from the Metropolitan Atlanta Congenital Defects Program (MACDP), a population-based birth defects surveillance system from 1998 - 2005. Children with CHD were defined by the nomenclature developed from the International Paediatric and Congenital Cardiac Code, an initiative developed by members of the Society of Thoracic Surgeons (STS) and the European Association for Cardio-Thoracic Surgery.^8^ All identified cases were limited to surgical diagnoses used by the STS congenital heart surgery database. This study by Reller et al reports an overall CHD prevalence of 8.14 per 1,000 live births.^3^

Population-based empirical data on the epidemiology and prevalence of children with CHD have been limited. Marelli and colleagues report contemporary prevalence estimates of CHD using Canadian health services databases.^9^ Infants were identified by administrative diagnosis and surgical procedural codes, reporting an observed prevalence rate of 8.21 per 1,000 from 1983 - 2010. A more recent model developed from the Global Burden of Diseases, Injuries and Risk Factors Study found a 4.2% increase in global CHD prevalence since 1990.^10^ We continue to lack robust and empirical U.S.-based population estimates.^9^

Advancements in medical care and surgical treatments have significantly increased the survival of CHD individuals.^11–13^ Despite such progress, research continues to show that children with CHD are associated with both the highest mortality and the highest inpatient resource use in pediatric care.^14^ In 2004, congenital abnormalities accounted more than 139,000 hospitalizations, of which over 30% were for cardiac and circulatory anomalies.^15^ In 2009, hospital admissions for CHD cost an estimated $5.6 billion annually, comprising 15% of costs for all pediatric care, while only accounting for 3.7% of pediatric admissions.^14, 16, 17^ It is now widely recognized that CHD is associated with frequent health care use and lifelong comorbidity.^9, 12^ The impact of ongoing disease burden includes depression, learning disabilities, asthma, cancer, obesity, rheumatologic disease, hypertension, stroke, and heart failure. The true magnitude of comorbidity burden is not known.^16, 18^

Estimating prevalence of children with CHD by healthcare administrative claims data presents a novel opportunity to study longitudinal patterns of this prominent disease. All-payer claims databases (APCDs) cache public and private claims for health care services provided to insured individuals within a state.^19^ Twenty-one states have established APCDs, and many others are currently in development.^20, 21^ The governance of APCDs, their funding, and the authority of APCDs to require claims submissions varies by state.

In the current study we present contemporary, longitudinal, population-based estimates for children with CHD. High-quality, population-based prevalence estimates of CHD prevalence is critical to support the understanding of pediatric CHD epidemiology, as it may lead to better insight into its etiology and care practices.^10^ A comprehensive assessment of the CHD health burden can support appropriate allocation of resources for diagnosis, care coordination, care access and cost-effective treatment strategies.

## METHODS

### DATA SOURCE: CO APCD

The Center for Improving Value in Health Care (CIVHC) is a nonprofit authorized by the state to collect Colorado (CO) claims data. The APCD includes medical claims and dates of service from commercial health plans, Medicare and Colorado’s Medicaid program. CO APCD includes hospital, clinic and claims files related to diagnosis, procedure, enrollment, member and provider data.

The current study is a retrospective study of children with CHD represented in the CO APCD. This is a longitudinal dataset spanning eight calendar years (January 1, 2012-December 31, 2019). Every individual was assigned a unique encrypted identifier on their first healthcare encounter and followed across all diagnoses, encounters, settings and years.

### STUDY POPULATION

Our study population included all children aged 18 years or younger on Jan 1, 2012, with a documented CO zip-code and at least one healthcare encounter in CO (N= 1,535,816) during the study period. Individuals with CHD were identified by the American Heart Association-American College of Cardiology (AHA-ACC) harmonized cardiac codes.^10^ Using this algorithm, CHD is defined by the *International Classification of Diseases, Ninth Revision,* or *International Statistical Classification of Diseases and Health Related Problems, Tenth Revision, (ICD-9/10)* in the primary diagnosis field (ICD-9: 745.xx, 746.xx, 747.xx; ICD-10: Q20.x, Q21.x, Q22.x, Q23.x, Q24.x, Q25.x, Q26.x, Q27.x, Q28.x) (Appendix Table 1). Our study population included well-recognized chromosomal abnormalities that are commonly associated with CHD. Children with co-occurring genetic abnormalities have often been excluded in CHD prevalence studies due to their more complex disease. These genetic abnormalities include Down syndrome, Turner syndrome, DiGeorge syndrome, trisomy 13, trisomy 18, Alagille syndrome, Noonan syndrome, Holt-Oram syndrome, Williams syndrome, Kabuki syndrome, VATER/VACTERL association, CHARGE association, Jacobsen syndrome and Williams-Beuren syndrome.^22–24^

**Table 1.** Characteristics of children with and without congenital heart disease in Colorado, 2012 – 2019.

|  | CHD<br>N= 30,512 (%) | NON-CHD<br>N = 1,565,815 (%) | P value |
| --- | --- | --- | --- |
| Age<sup>1</sup> |  |  |  |
| Newborn | 12,527 (41.1) | 301,746 (19.7) |  |
| Infant | 4,647 (15.2) | 90,632 (5.9) |  |
| Child | 7,238 (23.7) | 591,138 (38.5) |  |
| Youth | 6,100 (20.0) | 552,299 (36.0) | <0.001 |
| Sex<sup>2</sup> |  |  |  |
| Male | 15,843 (51.9) | 767,356 (49.9) | <0.001 |
| Primary insurer |  |  |  |
| Medicaid | 21,915 (71.8) | 897,634 (58.5) |  |
| Commercial | 8,597 (28.2) | 638,181 (41.6) | <0.001 |
| Rurality<sup>3</sup> |  |  |  |
| Urban | 26,957 (88.4) | 1,329,187 (86.6) | <0.001 |
| Region |  |  |  |
| Eastern | 947 (3.1) | 45,314 (3.0) |  |
| Front Range | 21,827 (71.5) | 1,055,938 (68.8) |  |
| Mountain | 5,148 (16.9) | 286,072 (18.6) |  |
| Southern | 1,427 (4.7) | 65,602 (4.3) |  |
| Western | 1,163 (3.8) | 82,889 (5.4) | <0.001 |
| Median household income<sup>4</sup> |  |  |  |
| Q1 | 6,965 (22.8) | 312,453 (20.3) |  |
| Q2 | 5,579 (18.3) | 268,015 (17.5) |  |
| Q3 | 6,178 (20.3) | 304,892 (19.9) |  |
| Q4 | 6,035 (19.8) | 316,775 (20.6) |  |
| Q5 | 5,755 (18.9) | 333,680 (21.7) | <0.001 |
| Diagnosis severity category<sup>5</sup> |  |  |  |
| Simple | 19,959 (65.4) |  |  |
| Moderately Complex | 6,068 (19.9) |  |  |
| Complex | 2,618 (8.6) |  |  |
| Other | 1,867 (6.1) | N/A |  |
| Complex chronic conditions<sup>6</sup> | 16,878 (55.3) | N/A |  |
| Genetic abnormalities<sup>7</sup> | 3,023 (10.0) | N/A |  |
| Disability<sup>8</sup> | 11,749 (38.5) | N/A |  |

1. Age at first encounter categorized as: Newborn (0 – 30 days old); Infant (31 – 365 days old); Child (366 days – 10 years old); Youth (>10– 18 years old) 2. Gender categorized as male or female. 3. Rurality defined by the 2010 Rural-Urban Commuting Area Codes 4. Median household income defined by the American Community Survey (2019) U.S. Census. 1<sup>st</sup> Quintile: \$24,427 – 51,461; 2<sup>nd</sup> Quintile: \$51,484 – 63,318; 3<sup>rd</sup> Quintile: \$63,745 – 74,648; 4<sup>th</sup> Quintile: \$74,809 – 94,875; 5<sup>th</sup> Quintile: \$95,050 – 147,421. 5. Severity category defined by ACHD Anatomy and Physiology Classification 6. Coexisting chronic conditions defined by the Pediatric Complex Chronic Conditions (PCCC) algorithm 7. Genetic abnormalities: Down syndrome, trisomy 13, trisomy 18, Turner syndrome, DiGeorge syndrome, Alagille syndrome, Noonan syndrome, Holt-Oram syndrome, Williams syndrome, Kabuki syndrome, VATER/VACTERL association, CHARGE association, Jacobsen syndrome, and Williams-Beuren syndrome 8. Disabilities defined by the Children with Disabilities Algorithm (CWDA)

Our cohort did not include diagnoses related to arrhythmias, cardiomyopathies, or transplants. We also excluded diagnoses that are specifically associated with adult disease as the particular anatomy and progression of disease for children is distinctively unique from adult diagnoses. (Appendix Table 1). To increase the specificity of the algorithm for our study population, manual audits of CHD diagnoses (N = 120) were independently adjudicated by two pediatric cardiologists specializing in congenital heart disease. The manual audit had 95% inter-rater agreement. Discrepancies in diagnoses were agreed upon through expert consensus.

### Comorbidities and Disabilities

Individual comorbidities were defined by the Pediatric Complex Chronic Conditions (CCC) V2 classification system. CCCs are defined as “any medical condition that can be reasonably expected to last at least 12 months (unless death intervenes) and to involve either several different organ systems or 1 organ system severely enough to require specialty pediatric care and probably some period of hospitalization in a tertiary care center”. A comprehensive set of ICD-9-CM and ICD-10-CM codes were identified as CCC and subsequently bracketed into 12 categories (cardiovascular, premature and neonatal, neurologic or neuromuscular, metabolic, gastrointestinal, renal, respiratory, hematologic or immunologic, malignancy, technology dependency, transplantation, and other congenital or genetic condition).^25^ In our evaluation, we omitted categories related to cardiovascular and other congenital or genetic complex conditions as those are known to occur simultaneously with CHD.

Individual disabilities were defined by the Children with Disabilities Algorithm (CWDA), a validated, claims-based tool for identifying children with disabilities.^26^ CWDA classifies children as having a disability as those “who have long-term physical, mental, intellectual, or sensory impairments which in interaction with various barriers may hinder their full and effective participation in society on an equal basis with others.” CWDA also accounts for intellectual and mental conditions that “limit children’s ability to fully participate in society”. There are 17 distinct categories within the CWDA based on the disability concepts and definitions articulated by the World Health Organization (WHO).

### Spatial attributes

Individuals were categorized as urban-or rural residing by joining home zip-codes to 2010 Rural-Urban Commuting Area (RUCA) categorizations.^27^ RUCA codes categorize U.S. census tracts on measures of population density, urbanization, and daily commuting. RUCA codes used in this study were based on data from the 2010 decennial census. Rural is defined as “towns with populations below 10,000 and surrounding commuter areas with more than one-hour driving distance to the closest city”.^27 28^ Median household income estimates were derived from American Community Survey (ACS), a publicly available U.S. Census Bureau survey that enables linkage of community characteristics with individual zip-code tabulation area (ZCTA) codes.^29^

### Severity categories

CHD severity categories were defined by the American College of Cardiology/ American Heart Association guidelines for the Management of Adults with Congenital Heart Disease (ACHD). These guidelines define CHD severity within an anatomic and physiological framework.^30^ Category designations are clinically relevant and structured to facilitate incorporation of administrative data to inform prevalence levels and trends. These three severity categories are descriptively named as: simple, moderately complex and complex (Appendix Table 2). We included an additional fourth category ‘other’ to encompass remaining, uncategorized diagnoses. For individuals that had multiple diagnoses across different severity categories, we ranked codes hierarchically so that the most severe lesion would be assigned with the primary diagnosis (i.e. ‘complex’ diagnosis of transposition of the great vessels diagnosis would be considered primary even if child also had a ‘simple’ septal defect). We further assigned CHD diagnoses into anatomically oriented and clinically relevant lesion categories (Appendix Table 3).

**Table 2.** Most common diagnoses for children with congenital heart disease, by lesion category.

| **Lesion Category** | **N (%)** |
| --- | --- |
| Atrial septal defect | 24.4 |
| Ventricular septal defect | 11.3 |
| Patent ductus arteriosus | 10.6 |
| Aortic insufficiency | 10.0 |
| Pulmonary artery abnormality | 5.9 |
| Other congenital anomalies of the heart | 18.2 |

**Table 3.**
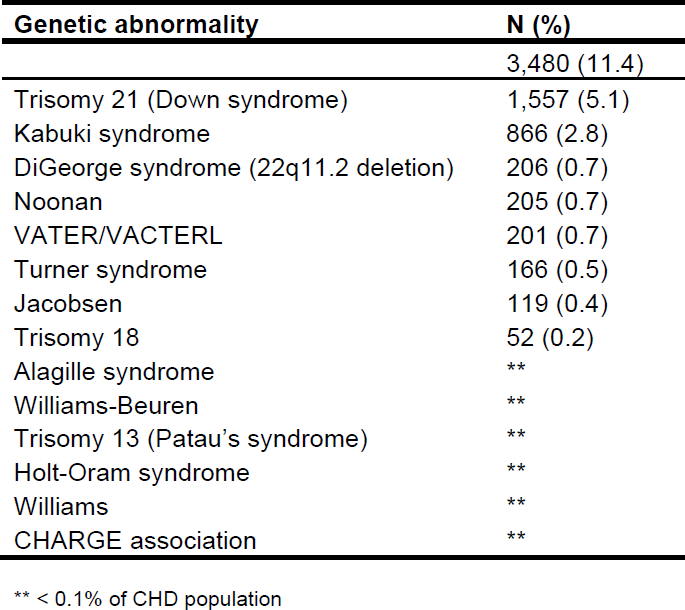
Prevalence of children with congenital heart disease in Colorado that have a concurrent genetic abnormality. N = 3.480 (11%); 405 (with more than 1 genetic diagnosis)

CHD diagnoses were analyzed by sociodemographic and geographic factors. Age at the time of first encounter was categorized as newborn (0 – 30 days), infant (31 – 365 days), child (366 – 10 years old), and youth (>10 – 18 years old). Sex was characterized as a dichotomous variable, with male as the referent. Primary insurance provider was assigned to the insurer associated with the most claims by individual and categorized by commercial/self-pay or Medicaid. Median household income was categorized by quintile (lowest quintile: $24,427 – 51,461; highest quintile: $95,050 – 147,421).

### Statistical Analysis

Prevalence is defined as the proportion of individuals in a population who have a disease at a specific point in time.^31^. We calculated the observed prevalence of children with CHD as the number of patients diagnosed with CHD in a given year (numerator), divided by the CO pediatric population in that year (denominator). Prevalence was reported as the number of unique CHD diagnosed children per 1,000 individuals. We applied the Cochran-Mantel-Haenszel test and Cochran-Armitage test for nonzero correlation to test for yearly trends. Categorical variables are described as frequencies and proportions and compared through chi^2^ tests.

We used direct standardization to calculate adjusted prevalence rates across the study period. The direct method of adjusting for differences among populations involves computing the overall rates that would result if all populations had the same frequency distribution. Direct standardization adjusts for any distortion caused by different distributions between the general population and study population. In the initial model, we adjusted for age, sex, primary insurance provider, region of residence, urban-rural residence and median household income. We performed tests of collinearity to refine our model. In the final model, we adjusted for age, sex, primary insurance provider and urban-rural residence.

## RESULTS

Figure 1. illustrates the derivation of the 1,565,387 children in the CO APCD health claims database during our longitudinal study period. We identified 30,512 unique children with CHD (received a CHD diagnosis at any point during the study period), comprising 1.9% (95% CI: 1.93 – 1.97) of the pediatric population.

**Figure 1.**
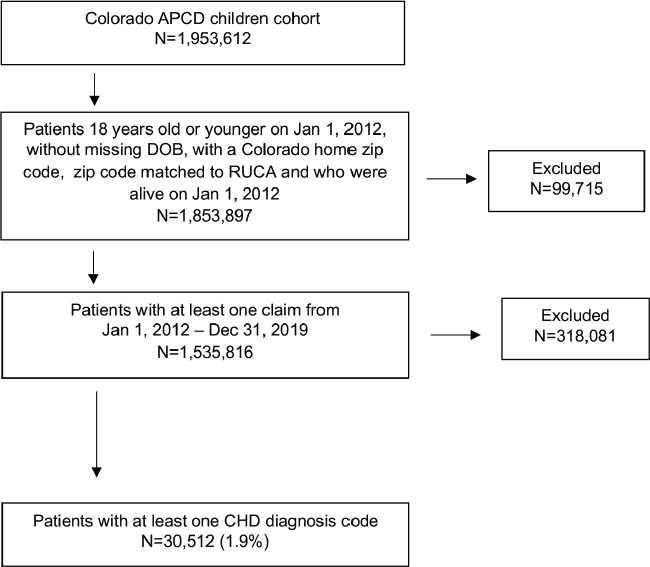
Derivation of cohort. Flow diagram of study subjects.

Prevalence by demographics, insurance provider, disease severity and community characteristics are summarized in Table 1. In total, 48.1% were female, 56.3% had their first CHD encounter before the age of one, 88.4% resided in urban core areas, and 71.8% were primarily insured by Medicaid.

Most children with CHD were diagnosed with simple, non-complex cardiac lesions (65.4%) and 19.9% were diagnosed with a moderately complex CHD lesion. By anatomically oriented category, atrial septal defect was most commonly diagnosed, comprising nearly 25% of all diagnoses, ventricular septal defects accounted for 11% and patent ductus arterioses were diagnosed in 11% of all diagnoses (Table 2).

Among the children with CHD, 3,023 (11%) were diagnosed with a genetic disorder, and 405 (1.3%) had more than one genetic diagnosis. Trisomy 21 (Down’s syndrome) was diagnosed in 1,557 children (5.1%) and 866 (2.8%) had Kabuki syndrome. Other genetic disorders frequently associated with CHD in our cohort included DiGeorge syndrome, Noonan syndrome, VATER/VACTERL, Turner syndrome, Jacobsen syndrome and Trisomy 18 (Table 3).

More than half of the children with CHD also experienced at least one concurrent chronic complex condition (55.3%), as identified by the CCC algorithm. Among those with CCCs, 7,626 (45.2%) had multiple CCC diagnoses in addition to CHD. The most common chronic complex conditions were related to respiratory, metabolic, gastrointestinal, neurologic and neuromuscular conditions. (Table 4).

**Table 4.** Prevalence of children with congenital heart disease that have concurrent complex chronic conditions.

In our CHD cohort, 11,749 (38.5%) received at least one disability diagnosis during the study period, as defined by the CWDA. There were 5,484 (18.0%) children with CHD that were treated for more than one disability. Nervous system and mental health disorders were most common (23.1% and 16.7%, respectively). We also found that children with CHD were treated for respiratory, circulatory, digestive, and genitourinary diseases. (Table 5).

**Table 5.** Prevalence of children with congenital heart disease that have concurrent disability.

| **Disability** | **N (%)** |
| --- | --- |
|  | 11,749 (38.5) |
| Diseases of the Nervous System and Sense Organs | 7,040 (23.1) |
| Mental Health Disorders | 5,085 (16.7) |
| Diseases of the Respiratory System | 3,361 (11.0) |
| Congenital Anomalies | 3,357 (11.0) |
| Certain Conditions Originating in the Perinatal Period | 2,514 (8.2) |
| Diseases of the Circulatory System | 1,873 (6.1) |
| Diseases of the Digestive System | 1,863 (6.1) |
| Diseases of the Genitourinary System | 1,393 (4.6) |
| Supplemental Classification of Factors Influencing Health Status | 687 (2.3) |
| Systems, Signs and Ill-Defined Conditions | ** |
| Endocrine, Nutritional and Metabolic Diseases, and Immunity Disorders | ** |
| Injury and Poisoning | ** |
| Neoplasms | ** |
| Infectious and Parasitic Diseases | ** |

\*\* < 1%

### Geography

Figures 2 and 3 illustrate the population density of all CO residing children and children with CHD in our study, respectively. While we observe a higher density of non-CHD children residing in the western and mountain regions, we note strong similarities between both cohorts in population clusters along the Front Range region.

**Figure 2.**
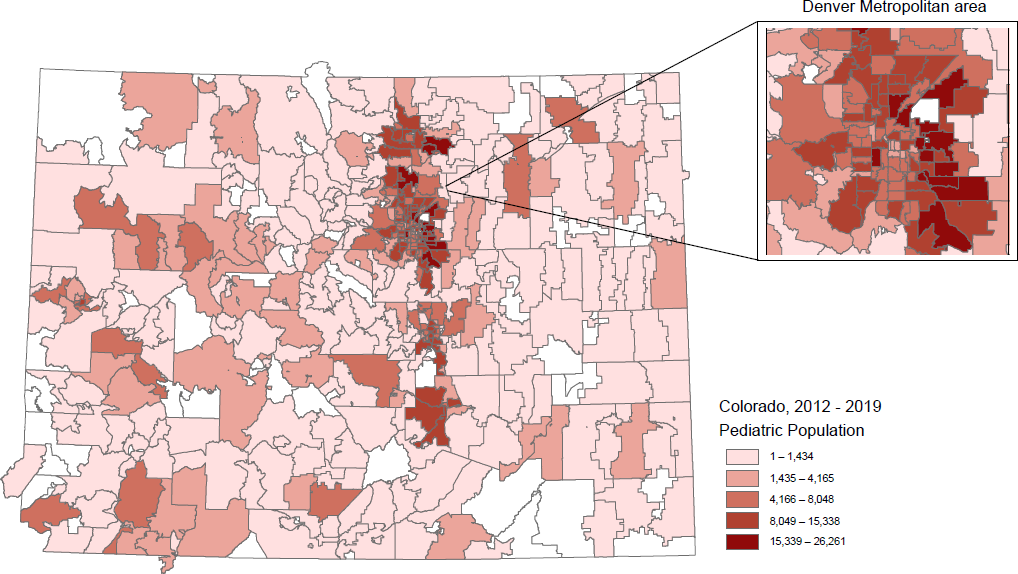
Population density map of children residing in CO, 2012 – 2019. The population density map illustrates the aggregate CO pediatric population at the zip-code tabulation area (ZCTA) level. The population is further categorized by quintile.

## DISCUSSION

We present the first U.S. population-based study to longitudinally estimate the prevalence and comorbidity burden of children diagnosed with CHD in the U.S. During 2012 to 2019, there were 30,512 children identified with CHD in CO, comprising 1.9% of the pediatric population. In comparison with previously reported estimates, our higher reported prevalence may be explained by the inclusion of children with concurrent genetic abnormalities, which have not been previously accounted for, as well as diagnoses captured after infancy. Our study summarizes the significant and complex health burden that children with CHD frequently experience, with more than half having at least one comorbid condition and 45% have multiple.

Most children with CHD were diagnosed with simple, non-complex cardiac lesions, and less than one-fifth of the children were diagnosed with a moderately complex lesion. There was an increased prevalence in ventricular and atrial septal defect subcategories, possibly related to improved detection or diagnosis after infancy. Ventricular septal defects are the most common form of CHD and studies show that at least 25% of all ventricular septal defects spontaneously close.^32^ Retaining an initial diagnosis of ventricular or atrial septal defect across the study period may also explain the increased prevalence.

CHD occurs in association with other anomalies or as part of an identified syndrome in 25 – 40% cases.^33^ We found that 11% of children with CHD were also diagnosed with a genetic abnormality. It has been reported that fifty-percent of individuals born with Trisomy 21 (Down’s syndrome) also have atrial and ventricular septal defects, and approximately one-third of females with Turner syndrome have CHD.^23^ In our study, we report a similar proportion of children diagnosed with both CHD and Trisomy 21.

Improved survival of children with CHD has led to increased attention to extracardiac complex conditions, often found across multiple body systems.^16, 34–36^ We found that over half of the children diagnosed with CHD have at least one coexisting chronic complex condition and 2 in 5 children have a disability in addition to CHD. The most common comorbidities were respiratory, metabolic and neurologic conditions but a significant proportion of children also had diagnoses related to renal, immune and malignant conditions.

It has been reported that long-term neurodevelopmental and cognitive disabilities are the most common comorbidities for children with CHD.^37, 38^ There are multiple theories to potentially explain the high prevalence of mental health disorders seen in this subset of children, such as abnormalities of in utero brain development and metabolism resulting in more saturated cerebral blood flow and underlying genetic syndromes, parental mental health disorders, prematurity, cardiac bypass surgery, and prolonged hospitalizations are posited as factors that could increase the risk of neurodevelopmental disabilities.^39–42^ A study of youths with CHD reported 7 times higher odds of anxiety and attention deficit/hyperactivity disorder (ADHD), compared to those without CHD.^39^ Applying validated algorithms, we found that over 25% of children with CHD also experience mental health disability. To date, we are the first to report the prevalence of mental health disabilities for children with CHD, both longitudinally and across healthcare settings.

Most studies elect to exclude children with chromosomal syndromes and genetic conditions, as it has been argued that this subset of children are uniquely complex. ^43, 44^ Proportion estimates of CHDs associated with a chromosomal abnormality vary widely from 9% to 18%, depending on study location, inclusion criteria for children with CHDs, and overall study population.^45^ We are the first to report on the prevalence of other chromosomal abnormalities such as 22q11.2 deletion (DiGeorge Syndrome), tristomy 21 (Down syndrome), tristomy 18 and tristomy 13, that historically have been excluded in population estimates.^45^

Our analysis has several notable advances compared with previous prevalence estimates. First, the prevalence analysis represent CHD children at the time of their first healthcare encounter, not at the time of their first CHD diagnosis. This approach allowed us to evaluate CHD longitudinally and include children who may have been diagnosed before the study or those who present after infancy.^10, 46, 47^ Many previously published studies identified children with CHD by active case-finding, or simply limited the population to newborns and infants. These methodologies may result in underestimation of less severe lesions more likely to manifest after the first year of life.^48, 49^ Second, we included all CHD diagnoses – regardless of setting, lesion severity, or surgical intervention. Many frequently cited CHD prevalence studies restricted analyses to only simple lesions, only severe lesions, or only CHD diagnoses requiring surgery.^1, 3, 6, 9, 49, 50^ Third, we incorporated many new sources of data and applied validated algorithms to estimate comorbidity burden. Finally, a fundamental strength of our data was the ability to longitudinally include children of all insurance types, throughout eight consecutive years.

The present study has several limitations. First, identification of children with CHD was made by administrative diagnoses codes ICD-9-CM and ICD-10-CM, thus both inadvertently excluding and including patients who were either misdiagnosed or miscoded. In addition, administrative codes often lack greater specificity compared with clinical nomenclature.^38^ Second, and relatedly, although subtyping of CHD is expanded in our dataset, it has yet to capture the full spectrum of disease. This is particularly true for defects that have a wide variety of presentations and severities. Third, we did not modify our analysis based on assumptions of spontaneous regression or surgical repair (iatrogenic regression). While children with small or simple CHD lesions may not require immediate or lifelong treatment, studies have shown small lesions are commonly associated with health complications requiring lifelong care.^1, 51–53^

## CONCLUSION

Knowledge of prevalence patterns of CHDs is important for improving our understanding of the magnitude of the disease, in both the population and extent of care necessary for children with CHD. An accurate assessment of the absolute and relative burden of CHD, as well as related comorbidities, on a population-based scale will support appropriate allocation of resources for diagnosis, care coordination, care access and cost-effective treatment strategies.

## Data Availability

Data is available through the Colorado Center for Improving Value in Health Care (CIVHC)

## Acknowledgments

None

---

**Supplementary Table 1.** CHD diagnoses and ICD-9/10 Mappings.

| ICD9 | ICD9 DX Description | ICD10 | ICD10 DX Description |
| --- | --- | --- | --- |
| 394 | Diseases of mitral valve | I34.0 | Nonrheumatic mitral (valve) insufficiency |
| 395 | Diseases of aortic valve | I34.8 | Other nonrheumatic mitral valve disorders |
| 396.0 | Mitral/aortic stenosis | I37.0 | Nonrheumatic pulmonary valve stenosis |
| 424.0 | Mitral valve disorders | I37.8 | Other nonrheumatic pulmonary valve disorders |
| 424.0 | Mitral valve disorders | Q21.3 | Tetralogy of fallot |
| 424.3 | Pulmonary valve disorders | Q28.8 | Other specified congenital malformations of circulatory system |
| 745.2 | Tetralogy of fallot | Q20.0 | Common arterial trunk |
| 746 | Congenital anomalies of heart. | Q20.3 | Discordant ventriculoarterial connection |
| 747.89 | Circulatory anomaly NEC | Q20.1 | Double outlet right ventricle |
| 745.0 | Common truncus | Q20.5 | Discordant atrioventricular connection |
| 745.10 | Complete transposition of great vessels | Q20.3 | Discordant ventriculoarterial connection |
| 745.11 | Double outlet right ventricle | Q20.8 | Other congenital malformations of cardiac chambers and connections |
| 745.12 | Corrected transposition of great vessels | Q20.4 | Double inlet ventricle |
| 745.19 | Other transposition of great vessels | Q21.0 | Ventricular septal defect |
| 745.19 | Other transposition of great vessels | Q21.1 | Atrial septal defect |
| 745.3 | Common ventricle | Q21.2 | Atrioventricular septal defect |
| 745.4 | Ventricular septal defect | Q21.2 | Atrioventricular septal defect |
| 745.5 | Ostium secundum type atrial septal defect | Q21.2 | Atrioventricular septal defect |
| 745.60 | Endocardial cushion defect, unspecified type | Q20.8 | Other congenital malformations of cardiac chambers and connections |
| 745.61 | Ostium primum defect | Q21.8 | Other congenital malformations of cardiac septa |
| 745.69 | Other endocardial cushion defects | Q21.9 | Congenital malformation of cardiac septum, unspecified |
| 745.8 | Other bulbus cordis anomalies and anomalies of cardiac septal closure | Q22.3 | Other congenital malformations of pulmonary valve |
| 745.8 | Other bulbus cordis anomalies and anomalies of cardiac septal closure | Q22.0 | Pulmonary valve atresia |
| 745.9 | Unspecified defect of septal closure | Q22.1 | Congenital pulmonary valve stenosis |
| 746.00 | Congenital pulmonary valve anomaly, unspecified | Q22.2 | Congenital pulmonary valve insufficiency |
| 746.01 | Atresia of pulmonary valve, congenital | Q22.9 | Congenital malformation of tricuspid valve, unspecified |
| 746.02 | Stenosis of pulmonary valve, congenital | Q22.5 | Ebstein's anomaly |
| 746.09 | Other congenital anomalies of pulmonary valve | Q23.0 | Congenital stenosis of aortic valve |
| 746.1 | Tricuspid atresia and stenosis, congenital | Q23.1 | Congenital insufficiency of aortic valve |
| 746.2 | Ebstein's anomaly | I34.2 | Nonrheumatic mitral (valve) stenosis |
| 746.3 | Congenital stenosis of aortic valve | Q23.3 | Congenital mitral insufficiency |
| 746.4 | Congenital insufficiency of aortic valve | Q23.4 | Hypoplastic left heart syndrome |
| 746.5 | Congenital mitral stenosis | Q24.4 | Congenital subaortic stenosis |
| 746.6 | Congenital mitral insufficiency | Q24.2 | Cor triatriatum |
| 746.7 | Hypoplastic left heart syndrome | Q24.3 | Pulmonary infundibular stenosis |
| 746.81 | Subaortic stenosis | Q24.8 | Other specified congenital malformations of heart |
| 746.82 | Cor triatriatum | Q24.5 | Malformation of coronary vessels |
| 746.83 | Infundibular pulmonic stenosis | Q24.0 | Dextrocardia |
| 746.84 | Obstructive anomalies of heart, not elsewhere classified | Q24.1 | Levocardia |
| 746.85 | Coronary artery anomaly | Q24.8 | Other specified congenital malformations of heart |
| 746.87 | Malposition of heart and cardiac apex | Q23.8 | Other congenital malformations of aortic and mitral valves |
| 746.87 | Malposition of heart and cardiac apex | Q24.8 | Other specified congenital malformations of heart |
| 746.87 | Malposition of heart and cardiac apex | Q20.9 | Congenital malformations of cardiac chambers and connections, unspecified |
| 746.89 | Other specified congenital anomalies of heart | Q24.9 | Congenital malformation of heart, unspecified |
| 746.89 | Other specified congenital anomalies of heart | Q25.0 | Patent ductus arteriosus |
| 746.9 | Unspecified congenital anomaly of heart | Q25.1 | Coarctation of aorta |
| 746.9 | Unspecified congenital anomaly of heart | Q25.40 | Congenital malformation of aorta unspecified |
| 747.0 | Patent ductus arteriosus | Q25.45 | Double aortic arch |
| 747.10 | Coarctation of aorta (preductal) (postductal) | Q25.46 | Tortuous aortic arch |
| 747.20 | Anomaly of aorta, unspecified | Q25.47 | Right aortic arch |
| 747.21 | Anomalies of aortic arch | Q25.29 | Other atresia of aorta |
| 747.21 | Anomalies of aortic arch | Q25.3 | Supravalvular aortic stenosis |
| 747.21 | Anomalies of aortic arch | Q25.41 | Absence and aplasia of aorta |
| 747.22 | Atresia and stenosis of aorta | Q25.42 | Hypoplasia of aorta |
| 747.22 | Atresia and stenosis of aorta | Q25.43 | Congenital aneurysm of aorta |
| 747.29 | Other anomalies of aorta | Q25.44 | Congenital dilation of aorta |
| 747.29 | Other anomalies of aorta | Q25.48 | Anomalous origin of subclavian artery |
| 747.29 | Other anomalies of aorta | Q25.49 | Other congenital malformations of aorta |
| 747.29 | Other anomalies of aorta | Q25.5 | Atresia of pulmonary artery |
| 747.29 | Other anomalies of aorta | Q25.71 | Coarctation of pulmonary artery |
| 747.29 | Other anomalies of aorta | Q25.72 | Congenital pulmonary arteriovenous malformation |
| 747.31 | Pulmonary artery coarctation and atresia | Q25.6 | Stenosis of pulmonary artery |
| 747.31 | Pulmonary artery coarctation and atresia | Q25.79 | Other congenital malformations of pulmonary artery |
| 747.32 | Pulmonary arteriovenous malformation | Q26.9 | Congenital malformation of great vein, unspecified |
| 747.39 | Other anomalies of pulmonary artery and pulmonary circulation | Q26.2 | Total anomalous pulmonary venous connection |
| 747.39 | Other anomalies of pulmonary artery and pulmonary circulation | Q26.3 | Partial anomalous pulmonary venous connection |
| 747.40 | Anomaly of great veins, unspecified | Q26.0 | Congenital stenosis of vena cava |
| 747.41 | Total anomalous pulmonary venous connection | Q26.1 | Persistent left superior vena |
| 747.42 | Partial anomalous pulmonary venous connection | Q26.8 | Other congenital malformations of great veins |
| 747.49 | Other anomalies of great veins | Q28.8 | Other specified congenital anomalies of circulatory system |
| 747.49 | Other anomalies of great veins | Z87.74 | Personal history of (corrected) congenital malformations of heart and circulatory system |
| 747.49 | Other anomalies of great veins | Q25.21 | Interruption of aortic arch |
| 747.89 | Other specified congenital anomalies of circulatory system |  |  |
| V13.65 | Personal history of (corrected) congenital malformations of heart and circulatory system |  |  |

**Table 2.** Disease Severity Categories and ICD-9/10 Mappings

| ICD9 | ICD-9 DESCRIPTION | ICD10 | ICD-10 DESCRIPTION |
| --- | --- | --- | --- |
| **SIMPLE** |  |  |  |
| 746.0 | Anomalies of pulmonary valve | Q22.1 | Congenital pulmonary valve stenosis |
| 746.00 | Pulmonary valve anomaly, unspecified | Q25.0 | Patent ductus arteriosus |
| 746.02 | Stenosis, congenital | Q21.0 | Ventricular septal defect |
| 747 | Patent ductus arteriosus | Q21.1 | Atrial septal defect |
| 745.4 | Ventricular septal defect | Q26.0 | Congenital stenosis of vena cava |
| 745.5 | Ostium secundum type atrial septal defect | Q26.8 | Other congenital malformations of great veins |
| 747.4 | Anomalies of great veins* | Q26.9 | Congenital malformation of great vein, unspecified |
| 747.40 | Anomaly of great veins, unspecified* |  |  |
| 747.49 | Other anomalies of great veins* |  |  |
| 424.3 | Pulmonary valve disorder |  |  |
| **MODERATELY COMPLEX** |  |  |  |
| 746.2 | Ebstein's anomaly | Q22.2 | Congenital pulmonary valve insufficiency |
| 746.5 | Congenital mitral stenosis | Q22.3 | Other congenital malformations of pulmonary valve |
| 746.6 | Congenital mitral insufficiency | Q22.5 | Ebstein's anomaly |
| 746.8 | Other specified anomalies of heart | Q22.8 | Other congenital malformations of tricuspid valve |
| 746.81 | Subaortic stenosis | Q22.9 | Congenital malformation of tricuspid valve, unspecified |
| 746.83 | Infundibular pulmonic stenosis | Q23.2 | Congenital mitral stenosis |
| 746.84 | Obstructive anomalies of heart, NEC | Q23.3 | Congenital mitral insufficiency |
| 747.1 | Coarctation of aorta | Q25.1 | Coarctation of aorta |
| 747.1 | Coarctation of aorta (preductal) (postductal) | Q25.3 | Supravalvular aortic stenosis |
| 745.2 | Tetralogy of Fallot | Q21.3 | Tetralogy of Fallot |
| 746.3 | Congenital stenosis of aortic valve | Q23.0 | Congenital stenosis of aortic valve |
| 746.4 | Congenital insufficiency of aortic valve | Q23.1 | Congenital insufficiency of aortic valve |
| 747.41 | Total anomalous pulmonary venous connection | Q26.2 | Total anomalous pulmonary venous connection |
| 745.6 | Endocardial cushion defects | Q21.2 | Atrioventricular septal defect |
| 745.60 | Endocardial cushion defect, unspecified type | Q21.4 | Aortopulmonary septal defect |
| 745.61 | Ostium primum defect | Q21.8 | Other congenital malformations of cardiac septa |
| 745.69 | Other AV canal* | Q21.9 | Congenital malformation of cardiac septum, unspecified |
| 745.8 | Other | Q26.3 | Partial anomalous pulmonary venous connection |
| 745.9 | Unspecified defect of septal closure | Q24 | Other congenital malformations of heart |
| 747.42 | Partial anomalous pulmonary venous connection | Q24.2 | Cor triatriatum |
| 746.82 | Cor triatriatum | Q24.3 | Pulmonary infundibular stenosis |
| 746.85 | Coronary artery anomaly | Q24.4 | Congenital subaortic stenosis |
| 746.86 | Congenital heart block | Q24.5 | Malformation of coronary vessels |
| 746.87 | Malposition of heart and cardiac apex | Q25.6 | Stenosis of pulmonary artery |
| 747.3 | Anomalies of pulmonary artery | Q25.7 | Other congenital malformations of pulmonary artery |
| 424 | Mitral valve disorder | Q25.71 | Coarctation of pulmonary artery |
|  |  | Q25.72 | Congenital pulmonary arteriovenous malformation |
|  |  | Q25.79 | Other congenital malformations of pulmonary artery |
|  |  | Q25.8 | Other congenital malformations of other great arteries |
|  |  | Q25.9 | Congenital malformation of great arteries, unspecified |
|  |  | Q26.4 | Anomalous pulmonary venous connection, unspecified |
| **COMPLEX** |  |  |  |
| 747.2 | Other anomalies of aorta | Q20.0 | Common arterial trunk |
| 747.20 | Anomaly of aorta, unspecified | Q20.1 | Double outlet right ventricle |
| 747.21 | Anomalies of aortic arch | Q20.2 | Double outlet left ventricle |
| 745.0 | Common truncus | Q20.3 | Discordant ventriculoarterial connection |
| 745.1 | Transposition of great vessels | Q22.0 | Pulmonary valve atresia |
| 745.10 | Complete transposition of great vessels | Q23.8 | Other congenital malformations of aortic and mitral valves |
| 745.11 | Double outlet right ventricle | Q23.9 | Congenital malformation of aortic and mitral valves, unspecified |
| 745.12 | Corrected transposition of great vessels | Q25.2 | Atresia of aorta |
| 745.19 | Transposition - Other* | Q20.4 | Double inlet ventricle |
| 746.01 | Atresia, congenital | Q22.4 | Congenital tricuspid stenosis |
| 745.3 | Common ventricle | Q22.6 | Hypoplastic right heart syndrome |
| 745.7 | Cor biloculare | Q23.4 | Hypoplastic left heart syndrome |
| 746.1 | Tricuspid atresia and stenosis, congenital | Q20.5 | Discordant atrioventricular connection |
| 746.7 | Hypoplastic left heart syndrome | Q20.6 | Isomerism of atrial appendages |
| 747.11 | Interruption of aortic arch | Q20.8 | Other congenital malformations of cardiac chambers and connections |
| 747.22 | Atresia and stenosis of aorta | Q20.9 | Congenital malformation of cardiac chambers and connections, unspecified |
|  |  | Q25.21 | Interruption of aortic arch |
|  |  | Q25.29 | Other atresia of aorta |
|  |  | Q25.40 | Congenital malformation of aorta unspecified |
|  |  | Q25.41 | Absence and aplasia of aorta |
|  |  | Q25.42 | Hypoplasia of aorta |
|  |  | Q25.49 | Other congenital malformations of aorta |
| **UNCATEGORIZED / OTHER** |  |  |  |
| 746.09 | Other | Q24.8 | Other specified congenital malformations of heart |
| 746.89 | Other | Q24.9 | Congenital malformation of heart, unspecified |
| 746.90 | Unspecified anomaly of heart | I34.2 | Eisenmenger's syndrome |
| 747.00 | Other congenital anomalies of circulatory system | Q24.1 | Levocardia |
|  |  | Q25.43 | Congenital aneurysm of aorta |
|  |  | Q25.44 | Congenital dilation of aorta |
|  |  | Q25.46 | Tortuous aortic arch |
|  |  | Q25.47 | Right aortic arch |
|  |  | Q25.48 | Anomalous origin of subclavian artery |

**Appendix Table 3.** Lesion categories and ICD-9/10 Mappings

|  | **Diagnosis** | **Codes** |
| --- | --- | --- |
| 1 | Non-specific mitral valve disease | 394, 424, I34.8 |
| 2 | Combined mitral and aortic disease | 396, Q23.8 |
| 3 | Mitral insufficiency | I34.0, 746.6, Q23.3 |
| 4 | Non-specific pulmonary valve disease | 424.3, 746.0, 746.09, I37.8, Q22.3 |
| 5 | Pulmonary stenosis | I37.0, 746.02, Q22.1 |
| 6 | Tetralogy of Fallot | 745.2, Q21.3 |
| 7 | Truncus arteriosus | 745, Q20.0 |
| 8 | Transposition of the great vessels | 745.10, 745.19, 745.3, Q20.3, Q20.4 |
| 9 | Other single ventricle disease (DORV, DILV) | 745.11, 745.3, Q20.1, Q20.4 |
| 10 | Corrected transposition of the great vessels | 745.12, Q20.5 |
| 11 | Ventricular septal defect | 745.4, 745.8, 745.9, Q21.0, Q21.8, Q21.9 |
| 12 | Atrial septal defect | 745.5, Q21.1 |
| 13 | Atrioventricular septal defect | 745.60, 745.61, 745.69, Q21.2 |
| 14 | Pulmonary atresia | 746.01, Q22.0, Q25.5, |
| 15 | Pulmonary insufficiency | Q22.2 |
| 16 | Tricuspid atresia/stenosis | 746.1, Q22.9 |
| 17 | Ebstein's anomaly | 746.2, Q22.5 |
| 18 | Aortic valve stenosis | 746.3, Q23.0 |
| 19 | Aortic insufficiency | 746.4, Q23.1 |
| 20 | Mitral stenosis | 746.5, I34.2 |
| 21 | Non-specific aortic valve disease | 395 |
| 22 | Hypoplastic left heart syndrome | 746.7, Q23.4 |
| 23 | Subaortic stenosis | 746.81, Q24.4 |
| 24 | Cor triatriatum | 746.82, Q24.2 |
| 25 | Pulmonary infundibular stenosis | 746.83, Q24.3 |
| 26 | Coronary artery anomaly | 746.85, Q24.5 |
| 27 | Patent ductus arteriosus | 747.0, Q25.0 |
| 28 | Coarctation of the aorta | 747.10, Q25.1 |
| 29 | Other aortic arch anomaly | 747.20, 747.21, 747.29, Q25.40, Q25.45, Q25.46, Q25.47, Q25.21, Q25.42, Q25.43, Q25.44, Q25.48, Q25.49 |
| 30 | Aortic atresia | 747.22, Q25.29, Q25.41 |
| 31 | Supravalvar aortic stenosis | Q25.3 |
| 32 | Pulmonary artery abnormality | 747.31, 747.32, 747.39, Q25.71, Q25.72, Q25.6, Q25.79 |
| 33 | Total anomalous pulmonary venous return | 747.41, Q26.2 |
| 34 | Partial anomalous pulmonary venous return | 747.42, Q26.3 |
| 35 | Other congenital anomalies of the heart | 746, 747.89, 746.84, 746.87, 746.89, 746.9, 747.40, 747.49, 747.89, V13.65, Q28.8, Q20.8, Q20.9, Q24.8, Q24.0, Q24.1, Q24.8, Q24.9, Q26.9, Q26.0, Q26.1, Q26.8, Q28.8, Z87.74 |

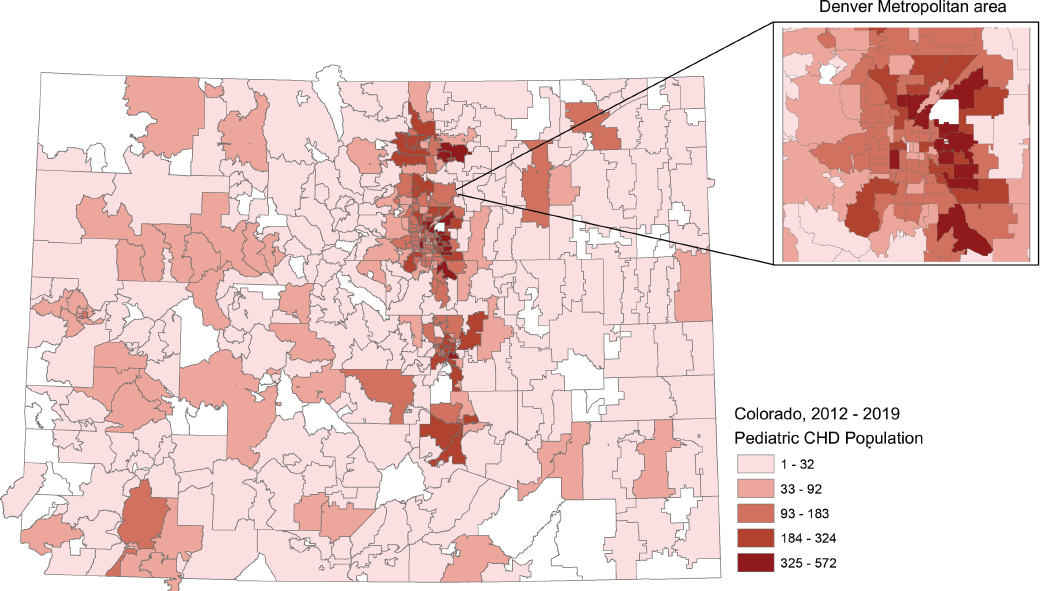

